# Effectiveness of asymptomatic patient screening for detection and control of multidrug-resistant organism transmission in healthcare settings – implications for infection control

**DOI:** 10.64898/2025.12.04.25341664

**Authors:** Christopher H. Connor, Claire L. Gorrie, Charlie K. Higgs, Torsten Seemann, Marcel Leroi, M. Lindsay Grayson, Benjamin P. Howden, Norelle L. Sherry, Jason C. Kwong

## Abstract

**Background:** Asymptomatic screening and contact precautions for multidrug-resistant organisms (MDRO) has been used to prevent transmission to other patients. However, the benefits are unclear due to differences in screening, challenges with accurately measuring transmission, and confounders when using clinical infections as an outcome measure. We aimed to compare the impact of different screening strategies on MDRO transmission among hospitalised patients using whole-genome sequencing (WGS) data.

**Methods:** We compared potential screening protocols targeting patients in i) intensive care, ii) high-MDRO-consequence wards, iii) high-MDRO-prevalence wards and iv) standard-risk wards from a 15-month prospective observational study of MDRO colonisation and infection. MDRO included *vanA* vancomycin-resistant *E. faecium*, extended-spectrum beta-lactamase (ESBL) producing *E. coli* and ESBL *K. pneumoniae*. WGS data from MDRO isolates were analysed with patient location data to define “probable”, “possible” and “unlikely” transmission events. The primary outcome was the number and proportion of probable transmission events identified with each screening strategy.

**Results:** 7475 patients were sampled with 668 MDRO cases (628 patients) identified from all samples (clinical samples plus asymptomatic screening). A strategy incorporating screening in ICU, high-consequence wards and high-prevalence wards detected significantly more MDRO cases (524/668 [78%] vs 120/668 [18%]; p<0.001) and probable transmission events (20/41 [49%] vs 5/41 [12%]; p<0.001) compared to MDRO surveillance based solely on clinical samples without screening. MDRO incidence (209.5 vs 78.8 cases per 1000 patients screened; RR 2.66; 95% CI 2.06–3.44; p<0.001) and transmission (44.7 vs 3.5 probable transmission events per 1000 patients screened; RR 12.77; 95% CI 5.97–27.0; p<0.001) was significantly higher on standard risk wards (no regular screening) than on high-consequence wards (weekly screening performed).

**Conclusion:** Active asymptomatic screening significantly enhanced the detection of target MDRO and MDRO transmission. Routine screening of patients with isolation of MDRO carriers in contact precautions was associated with lower rates of MDRO transmission.

**Key points:**

- MDRO surveillance based upon clinical infections alone missed the majority of MDRO incident cases and transmission events
- Asymptomatic screening for MDRO enhanced detection of cases and transmission
- MDRO transmission was lower on wards where asymptomatic screening was routinely performed

## INTRODUCTION

Healthcare associated infections (HAI) prolong patient admissions and negatively impact patient outcomes (1–3). These infections are further complicated by multidrug-resistant organisms (MDRO) where therapeutic options are limited (4). However, MDRO are adept at spreading within healthcare environments and can asymptomatically colonise patients (5).

Universal interventions such as hand hygiene and environmental cleaning are routinely employed but are unable to prevent all MDRO transmission. An active patient screening strategy for early case identification and isolation has been used particularly for high-consequence MDRO such as carbapenemase-producing Enterobacterales and *Candida auris* (6,7). Due to resource requirements, active screening strategies are often limited to “high-risk” groups such as room contacts of an index case and patients recently hospitalised in highly endemic areas. However, transmission and outbreaks of MDRO can still occur when MDRO-colonised cases are missed among non-isolated patients.

The optimal endpoint for studies assessing MDRO control measures is also uncertain. Previous studies have attempted to estimate or model the effectiveness of MDRO screening, but there are few transmission outcome data for screening strategies targeting MDRO that colonise the gut such as multidrug-resistant Enterobacterales and vancomycin-resistant enterococci (VRE) to inform these models due to the challenges in accurately capturing and identifying transmission (8,9).

Bacterial whole-genome sequencing (WGS) provides increased resolution to identify and evaluate potential MDRO transmission within hospitals (10) with recent progress towards implementing WGS for real-time detection of transmission (11). However, the ability of WGS to identify and assess MDRO transmission is dependent on MDRO screening practices.

In this study, we undertook a *post hoc* analysis of prospectively collected genomic surveillance data for MDRO in a single tertiary healthcare network to explore the impact of different screening strategies on MDRO transmission and control (11).

## METHODS

### Study population

The hospital network comprised three distinct facilities: one tertiary referral acute care centre and two subacute hospitals with a total of 770 inpatient care beds. Hospital wards accommodated 28–32 patient beds in a mixture of single and multi-occupancy rooms. Routine admission and thereafter weekly screening for MDRO was undertaken in several areas of the hospital – a) the Intensive Care Unit (ICU); b) “high-consequence” (immunocompromised patient) wards including Haematology, Oncology, bone marrow transplantation, and solid organ transplantation wards; and c) “high-prevalence” wards including Spinal Injury, Ventilatory Weaning, and Infectious Diseases wards that provided state-wide healthcare services and frequently accommodated patients repatriated from overseas healthcare facilities. Patients on high-prevalence wards were screened weekly in the context of high-consequence MDRO exposures and outbreaks. Other wards where asymptomatic screening was not routinely performed were considered “standard risk”. Patients on standard risk wards were occasionally screened for MDRO due to individual risk factors such as pre-transplant/chemotherapy assessment, planned transfer to high-consequence ward, recent overseas hospitalisation, or exposure to other reportable MDRO such as carbapenemase-producing organisms or *Candida auris* in accordance with state-wide guidelines. In addition, hospital-wide Point Prevalence Surveys (PPS) for MDRO colonisation involving screening of all admitted inpatients across the hospital network was performed biannually. During the 453-day study period, a PPS was conducted on four occasions. (Supplementary Table S1)

### Screening and isolation

Screening was undertaken by obtaining a rectal swab or faecal sample from admitted inpatients, as per routine clinical processes. Non-screening “clinical samples” comprised all other sample types that were obtained for the purpose of identifying clinical infections rather than screening for MDRO colonisation.

MDRO isolates were identified from screening and clinical samples and referred from the hospital network to a central ISO-accredited sequencing laboratory for whole-genome sequencing using standardised workflows as previously reported (11,12).

Patients identified with MDRO were placed into isolation under contact precautions, comprising the use of gowns and gloves in addition to hand hygiene and standard precautions, and placement in a single room. Shared patient equipment were cleaned and disinfected after each use in accordance with hospital protocols and rooms and bathrooms were cleaned upon patient discharge using a combined sodium hypochlorite-detergent product.

### Stratification

To determine the impact that varied screening practices have on transmission detection we compared potential screening strategies stratified into six tiers (Table 1 and Figure 1). For this study, three MDRO were included as indicator organisms – *vanA* vancomycin-resistant *Enterococcus faecium* (*vanA* VRE), extended-spectrum beta-lactamase (ESBL) phenotype *Klebsiella pneumoniae* (ESBL-Kp) and ESBL phenotype *Escherichia coli* (ESBL-Ec).

**Figure 1:**
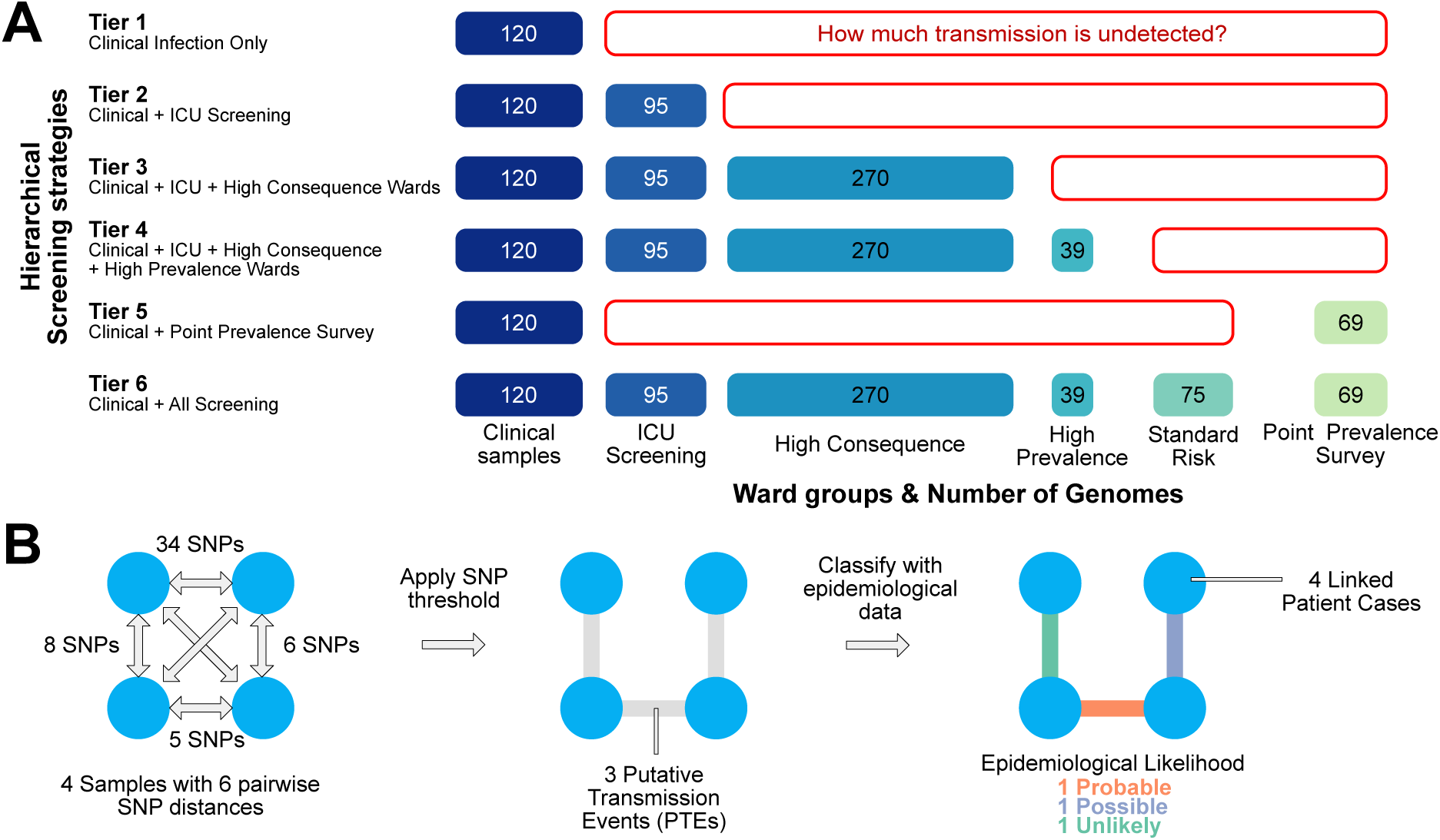
Diagram of screening strategy stratification and methodology for identifying and classifying Putative Transmission Events (PTE). Hierarchical stratification of active screening strategies and number of MDRO cases in each tier (A). Method for performing pairwise comparisons between all samples, identifying putative transmission events (PTE) by applying a SNP threshold and subsequently classifying PTE by incorporating epidemiological data (B).

**Table 1:** Stratification based on potential screening strategies.

| Tier | Isolates included |
| --- | --- |
| 1 | Clinical samples only |
| 2 | Clinical samples + screening in ICU only |
| 3 | Clinical samples + screening in ICU + high-consequence wards |
| 4 | Clinical samples + screening in ICU + high-consequence + high-prevalence wards |
| 5 | Clinical samples + point prevalence survey screening only |
| 6 | Clinical samples + all screening samples |

*ICU = Intensive Care Unit*
*High-consequence wards* included Haematology, Bone Marrow Transplantation, Oncology, Renal Transplantation, and Liver Transplantation wards.
*High-prevalence wards* included Spinal, Ventilation Weaning and Infectious Diseases wards.

### Measuring Transmission

Given that identification of a MDRO transmission event represents a hypothesised event based on epidemiological probability, we first defined Putative Transmission Events (PTE) based on genomic similarity using a previously validated approach (see Supplementary Materials) (13,14) To verify the likelihood of PTE representing actual transmission events, we used admission and patient bed movement data to classify events as previously described (11). Briefly, PTE were classified as ‘probable transmission’ if patients were on the same ward at the same time, ‘possible transmission’ if admitted to the same ward at a different time (within 60 days) or admitted to the same hospital at the same time; all other patient pairs were classified as ‘unlikely transmission’ (11).

### Outcomes

The primary outcome of interest was the number of probable transmission events identified with each screening strategy out of the total probable transmission events identified in the dataset. Other outcomes of interest were the number of MDRO cases detected, number of linked patients determined by the genomic similarity of their MDRO isolates, and number of patients sampled per detected case and probable transmission event for each screening strategy.

### Statistical analysis and data visualization

MDRO incidence (colonisation) was standardised to a rate per patients sampled, while MDRO infections were standardised to incidence rates per 1000 separations (completion of a patient episode of care e.g. discharge, transfer or death). Statistical tests of independence for categorical variables were performed using Fisher’s Exact Test and Chi-Square Tests, while comparisons of incidence and transmission rates were undertaken by calculating rate ratios using median-unbiased estimation (mid-p). Data were analysed, processed and visualized Python 3.12.4 and R v4.5.0.

## RESULTS

### Study population

Over a 15-month study period a total of 7,475 patients were sampled, yielding 668 cases of the target MDRO from 628 unique patients (Supplementary Figure S1). This comprised 489 ESBL-Ec, 98 ESBL-Kp and 81 *vanA* VRE cases (Figure 2, Table 2). The MDRO-colonised patient cohort had a median age of 66.5 years and comprised 61.0% male patients (Supplementary Figure S2). Of the 668 MDRO cases in the study period, 120 (18.0%) were identified from non-screening clinical samples obtained for suspected infections, and 548 (82.0%) through asymptomatic screening.

**Figure 2:**
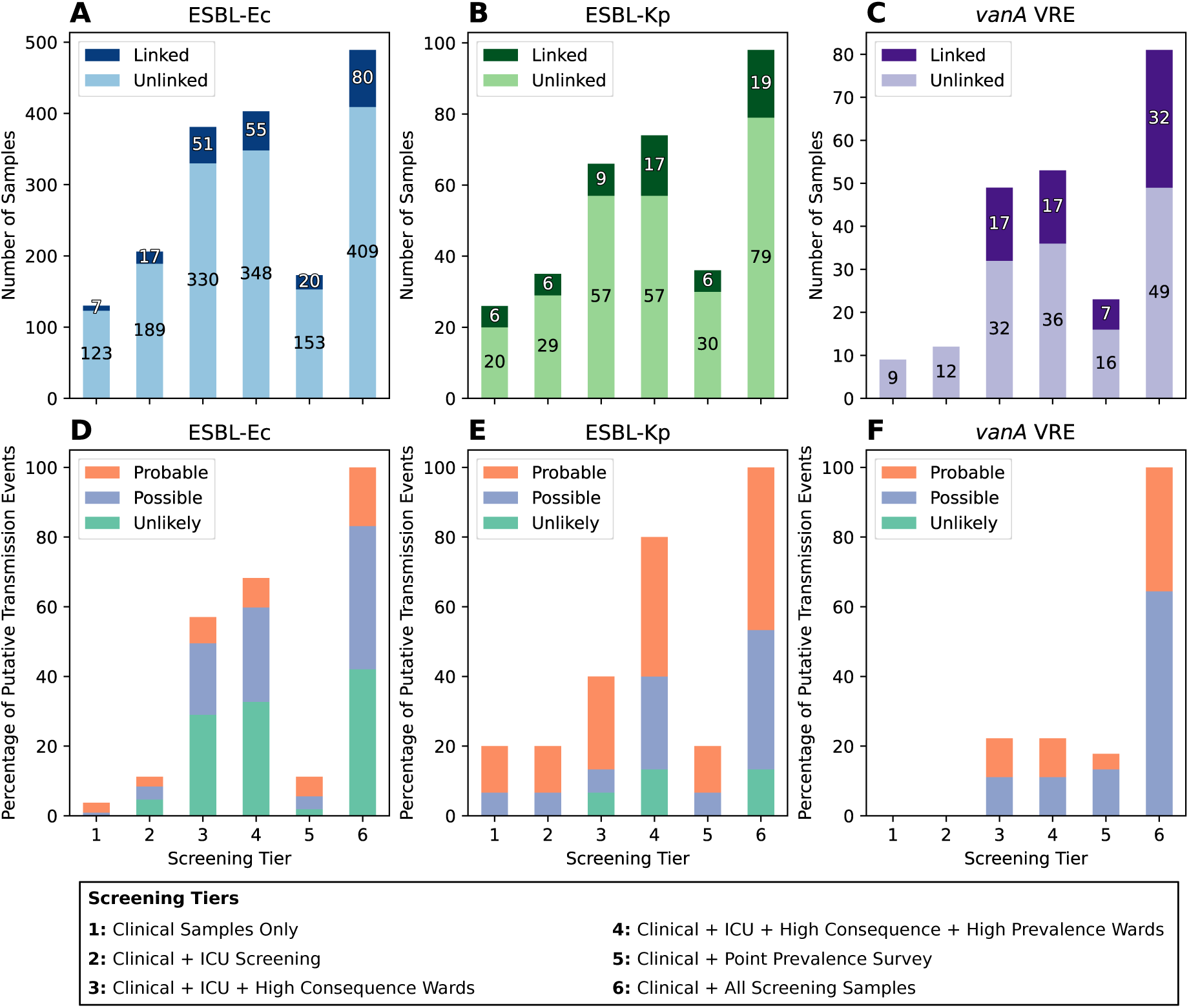
Number of patient cases and classification of Putative Transmission Events (PTE). Number of ‘Linked’ and ‘Unlinked’ cases at each screening tier (A – C). Linked cases are patient samples which have a PTE link to sample(s) from other patient(s), whilst unlinked cases were not within the SNP threshold of any others. Cases that are not genomically linked to another case are in a lighter colour, while cases that have a PTE to another case are in a darker colour. Case numbers for ESBL-Ec (A), ESBL-Kp (B) and *vanA* VRE (C). Epidemiological classification of PTEs (D – F). Patient bed movement data was used to classify PTE links into Probable, Possible or Unlikely. Epidemiological likelihood classification for ESBL-Ec (D), ESBL-Kp (E) and *vanA* VRE (F).

**Table 2:** MDRO cases by screening location.

| Sample Type/Location | Patients Screened | ESBL-Ec n (%) | ESBL-Kp n (%) | *vanA* VRE n (%) | Total n (%) | MDRO detections per 1000 patients screened |
| --- | --- | --- | --- | --- | --- | --- |
| Clinical samples – all areas | N/A | 98 (20%) | 18 (18%) | 4 (5%) | 120 (18%) | N/A |
| Screening – ICU | 1249 | 81 (17%) | 11 (11%) | 3 (4%) | 95 (14%) | 76.1 |
| Screening – High-consequence Wards | 3428 | 193 (39%) | 36 (37%) | 41 (51%) | 270 (40%) | 78.8 |
| Screening – High-prevalence Wards | 196 | 26 (5%) | 9 (9%) | 4 (5%) | 39 (6%) | 199.0 |
| Screening – Standard Risk Wards | 358 | 46 (9%) | 14 (14%) | 15 (19%) | 75 (11%) | 209.5 |
| PPS Screening – all areas | 2244 | 45 (9%) | 10 (10%) | 14 (17%) | 69 (10%) | 30.8 |
| **Total** | **7475** | **489** | **98** | **81** | **668** | **89.4** |

### MDRO incidence and clinical infections

The three indicator MDRO pathogens displayed differences in their distribution and yield from screening across the hospital (Table 2). Sampling occurred most frequently on high-consequence wards, which yielded 270 (40%) of the indicator MDRO cases. MDRO incidence rates on standard risk and high-prevalence wards were higher than on high-consequence wards. Overall incidence rates were 65.4 cases per 1000 patients sampled for ESBL-Ec, 13.1 cases per 1000 patients sampled for ESBL-Kp, and 10.8 cases per 1000 patients sampled for *vanA* VRE.

Fifty-seven of 548 (10%) cases detected through asymptomatic screening developed subsequent MDRO clinical infection during the study period, with collection of the clinical sample occurring a median of 18 days (IQR: 4–67 days) after the screening sample collection for these cases. The paired screening and clinical infection isolates were of the same sequence type and organism for 49/57 (86%) cases. A greater proportion of cases colonised with ESBL-Ec (20%) and ESBL-Kp (18%) developed clinical infections, compared to cases colonised with *vanA* VRE (5%). Clinical infection rates were highest in the ICU (31.25 per 1000 patient separations) and high-consequence wards (7.21 per 1000 patient separations), compared to standard risk wards (3.15 per 1000 patient separations) (Supplementary Table S2).

### Clinical samples missed the majority of new MDRO cases and transmission events

Using a surveillance scenario reliant upon cases identified from clinical samples, we identified only a minority of MDRO cases and very few genomically-linked patients using clinical infection isolates (Figure 2, Table 3). Only 13 of the total 131 (10%) genomically-linked MDRO cases were identified from clinical samples (7 ESBL-Ec, 6 ESBL-Kp), of which 5 represented probable transmission of the total 41 (12%) probable transmission events captured across the entire study.

**Table 3:** Genomically linked cases and Probable Putative Transmission Events by MDRO and screening location.

| **Linked Patient Cases** | **ESBL-Ec**<br>**n (%)** | **ESBL-Kp**<br>**n (%)** | ***vanA* VRE**<br>**n (%)** | **Total**<br>**n (%)** | **Linked Cases per 1000 patients**<br>**screened** |
| --- | --- | --- | --- | --- | --- |
| Clinical samples – all areas | 7 (9%) | 6 (32%) | 0 (0%) | 13 (10%) | N/A |
| Screening – ICU | 10 (13%) | 0 (0%) | 0 (0%) | 10 (8%) | 8.0 |
| Screening – High-consequence Wards | 34 (43%) | 3 (16%) | 17 (53%) | 54 (41%) | 15.8 |
| Screening – High-prevalence Wards | 4 (5%) | 8 (42%) | 0 (0%) | 12 (9%) | 61.2 |
| Screening – Standard Risk Wards | 12 (15%) | 2 (11%) | 8 (25%) | 22 (17%) | 61.5 |
| PPS Screening – all areas | 13 (16%) | 0 (0%) | 7 (22%) | 20 (15%) | 8.9 |
| **Total** | **80** | **19** | **32** | **131** | **17.5** |

| **Probable Putative Transmission Events** | **ESBL-Ec**<br>**n (%)** | **ESBL-Kp**<br>**n (%)** | ***vanA* VRE**<br>**n (%)** | **Total**<br>**n (%)** | **Probable Putative**<br>**Transmission Events per 1000**<br>**patients screened** |
| --- | --- | --- | --- | --- | --- |
| Clinical samples – all areas | 3 (17%) | 2 (29%) | 0 (0%) | 5 (12%) | N/A |
| Screening – ICU | 0 (0%) | 0 (0%) | 0 (0%) | 0 (0%) | 0.0 |
| Screening – High-consequence Wards | 5 (28%) | 2 (29%) | 5 (31%) | 12 (29%) | 3.5 |
| Screening – High-prevalence Wards | 1 (6%) | 2 (29%) | 0 (0%) | 3 (7%) | 15.3 |
| Screening – Standard Risk Wards | 6 (33%) | 1 (14%) | 9 (56%) | 16 (39%) | 44.7 |
| PPS Screening – all areas | 3 (17%) | 0 (0%) | 2 (13%) | 5 (12%) | 2.2 |
| **Total** | **18** | **7** | **16** | **41** | **5.5** |

### Active patient screening detected additional MDRO cases and hidden transmission events

In comparison, strategies that incorporated active screening of patients identified additional MDRO cases before they developed clinical infections. A strategy that incorporated active asymptomatic screening of ICU patients (Tier 2) in addition to identification of cases from clinical samples increased MDRO detection from 18% to 32% of all MDRO cases detected in the study. This corresponded to an additional 10 genomically-linked cases, but no new probable transmission events identified.

Increasing the number of patients screened resulted in incremental gains in capturing genomically-linked cases and probable transmission events (Figure 2 and Table 3). In comparison to no asymptomatic screening, an active surveillance strategy incorporating routine screening in ICU, high-consequence wards, and high-prevalence wards detected significantly more genomically-linked cases (89/131 [68%] vs 13/131 [10%]; p<0.001) and probable transmission events (20/41 [49%] vs 5/41 [12%]; p<0.001). As an alternative to routine weekly screening, conducting whole-of-hospital PPS every six months captured 25% of the linked cases and 10/41 (24%) probable transmission events.

MDRO transmission was highest on standard risk wards with 44.7 probable transmission events detected per 1000 patients screened (Table 3). In contrast to high-consequence wards where routine asymptomatic screening is performed weekly, MDRO transmission rates were significantly higher on standard risk wards (44.7 vs 3.5 probable transmission events per 1000 patients screened; RR 12.77; 95% CI 5.97–27.0; p<0.001). Although MDRO incidence on high-prevalence wards was similar to standard risk wards, fewer probable transmission events were detected (15.3 events per 1000 patients screened).

## DISCUSSION

MDRO pose a challenge to hospitals, adversely impacting patient outcomes and resulting in increased healthcare costs (15). Active patient screening allows hospitals to identify MDRO colonised patients and potentially implement controls such as isolation to reduce the risk of transmission to other patients, but there is little evidence on the optimal screening strategy in contexts where universal screening and/or isolation is not feasible (16–20). Here we demonstrate that active patient screening detects additional MDRO colonised patients that would otherwise have not been identified, with incremental gains for additional patients screened.

Previous studies have reported variable success with screening, with MDRO detection rates ranging between 27–87%(21,22). In our hospital, asymptomatic screening identified 465 of the 628 (74%) MDRO colonised or infected patients while the remainder were identified after developing clinical infections. Although impossible to know how many transmission events and additional MDRO cases were averted through early detection and isolation, the absence of asymptomatic screening would have missed detecting the majority of MDRO cases resulting in significantly more opportunities for transmission. The comparison between “standard risk” wards (asymptomatic screening not routinely undertaken) and “high-consequence” wards (patients screened weekly and isolated if MDRO are detected) is also notable. The MDRO incidence rate on “standard risk” wards was nearly three times the incidence on “high-consequence” wards, and had an associated transmission rate 14 times the rate on “high-consequence” wards, suggesting occult transmission of MDRO occurs frequently in the absence of targeted control measures such as case identification and isolation, and that those isolation measures including contact precautions were effective in reducing transmission.

Although several studies suggest that rates of MDRO-related healthcare-associated infection remain unaffected after discontinuing isolation practices for some MDRO such as VRE (23–25), there are are clear differences between MDRO and among different patient cohorts in their propensity for invasive infection following colonisation (26). The justification for screening and isolation in “high-consequence” patient cohorts (higher risk of invasive disease due to MDRO), and/or “high-prevalence” patient cohorts (higher risk of MDRO of significance such as carbapenemase-producing organisms and *Candidozyma auris*) in our institution follows this rationale. Furthermore, assessing the impact of controls such as screening and contact precautions with MDRO clinical infections as an outcome uses an indirect outcome measure, with multiple confounders influencing the transition from colonisation to clinical infection, and negates the ability to accurately measure the impact of interventions on preventing MDRO transmission.

Whole-genome sequencing (WGS) of bacterial isolates has been shown to enhance surveillance and outbreak investigations for MDRO (7,11) and is increasingly used for hospital infection prevention and control, but is reliant upon samples from identified cases (27,28). As shown in this study, sole reliance upon isolates from invasive infections (e.g. blood cultures) will miss the majority of cases required to infer transmission pathways to control outbreaks. Similarly, based on our data, an absence of MDRO transmission after discontinuing screening practices may actually represent missed detection of uncontrolled transmission. Despite the resource requirements for prospective WGS, recent studies have projected significant cost savings through integrated WGS surveillance (29). However as shown in our study, these cost savings can only be achieved through sufficient screening practices to identify MDRO cases and transmission events.

Some limitations should be noted. The data are derived from a cohort of patients in a single healthcare network which may limit extrapolation to other healthcare settings. However, comparing transmission is difficult without adjusting for confounding differences between healthcare facilities in screening and isolation practices and hospital infrastructure – factors difficult to control without large cluster-randomised crossover trials which are impractical for accurately. While previous studies have attempted to estimate broader impacts on population-level MDRO incidence (30), we focused on a smaller dataset with high-specificity definitions for transmission requiring both genomic and epidemiological criteria. Our reported rates of MDRO and transmission incidence are likely to underestimate the true rates due to imperfect sensitivity of MDRO screening and omission of cases detected outside our healthcare network, though these factors are likely to have been applied consistently across the study. Our data were limited to three indicator MDRO for pragmatic reasons, though we would anticipate similar findings for other multidrug-resistant Enterobacterales and enterococci. Additionally, our data report MDRO incidence in a setting where asymptomatic screening occurs regularly in many areas of the hospital and detected cases are isolated – actual numbers of MDRO cases may have been higher than in our scenarios if screening and isolation did not occur.

Our study provides real-world data to guide the relative gains from different screening approaches when implemented with isolation practices. Ultimately, the decision to employ a screening and isolation approach to MDRO will be influenced by the cost-benefit equation with different levels of benefit for different MDRO threats.

## Supporting information

Supplementary Accessions

## Abbreviations

ESBL: Extended-spectrum beta-lactamase
ICU: Intensive Care Unit
MDRO: Multidrug-resistant organism
PTE: Putative Transmission Event
SNP: Single-nucleotide polymorphism
VRE: Vancomycin-resistant *Enterococcus faecium*
WGS: Whole-genome sequencing

## Acknowledgements

Austin Health Department of Microbiology and Infection Prevention & Control team, and other members of the Controlling Superbugs Study Group.

## Data availability

Genome sequence data used in this study are available in the NCBI Sequence Read Archive (SRA) under BioProject PRJNA565795. A table of relevant sample IDs are provided in supplementary.

## Ethics

Ethics approval was obtained from the Melbourne Health Human Research Ethics Committee (HREC/13/MH/326) with local site approval provided by the Austin Health Office for Research.

