## Supplementary Material for "Effectiveness of asymptomatic patient screening for detection and control of multidrug-resistant organism transmission in healthcare settings – implications for infection control"

**Whole-genome sequencing**

Multi-drug resistant organism (MDRO) isolates were referred from the hospital network to a central ISO-accredited sequencing laboratory for whole-genome sequencing using standardised workflows (1). Genomic DNA was extracted from pure single colonies using a JANUS automated workstation (PerkinElmer) and Chemagic magnetic beads (PerkinElmer). DNA was sequenced using the Illumina NextSeq platform with Nextera XT libraries and protocols. Resulting reads were quality controlled using FastP v0.23.2 with the parameters cut front 3, sliding window size of 4, minimum quality 20 and minimum length of 36 (2).

**Identification of Putative Transmission Events (PTE)**

We defined Putative Transmission Events (PTE) based on genomic similarity of isolates using a previously validated approach for our analysis pipeline (3,4). The SNP distance between every pair of isolates (of the same MRDO) was calculated with SKA v1.0 (5). Sequencing reads in FASTQ format were used as input for SKA, along with a *k*-mer length of 19 and minor allele frequency of 0.01, as we have previously optimised (4). PTE were defined as SNP distances below a species-specific threshold, while the corresponding patients were referred to as “linked cases” (Figure 1 and S3). Thresholds were based on the within-patient genomic diversity observed in the study dataset as outlined below (4). Duplicate patient samples where the same MDRO and sequence type were identified were excluded from analysis. Patient samples were deduplicated at each analysis level, including only the earliest detection within the study period in each tier of analysis. For example, a patient who had a screening sample obtained in the ICU which identified a MDRO that preceded a clinical infection with that MDRO would have the clinical infection isolate included in the "Tier 1: Clinical samples only" analysis, but the screening sample would replace the clinical sample in the Tier 2, Tier 3, Tier 4 and Tier 6 analyses thereby simulating the earliest case detection under each surveillance strategy.

**Threshold Determination**

Species-specific thresholds to use for single linkage clustering were determined from the SNP distances observed between duplicate patient samples, as has been used previously (3). Isolates of the same MDRO and sequence type (ST) that were collected from the same patient within 90-days of each other were used to calculate SNP distances with SKA as described (Figure S4). The 90$^{th}$ quantile of this distribution was used as the SNP threshold for putative transmission events (PTE) identification. The resulting thresholds determined were 8 SNPs for ESBL-Ec, 12 SNPs for ESBL-Kp and 5 SNPs for *vanA* VRE.

| Hospital network | Hospital code | Hospital description | No. of inpatient beds[a] | High-risk wards | MDRO screening practices during study period and changes during study | Management of patients colonised or infection with MDRO |
|---|---|---|---|---|---|---|
| **A** | **A1** | Tertiary referral center, including ICU, solid-organ and haemopoietic stem cell transplant (HSCT) | 560 | ICU<br>Hematology/HSCT<br>Oncology<br>Renal Transplant<br>Liver Transplant<br>Spinal ward<br>Respiratory ward | **ESBL-Ec, ESBL-Kp and *vanA* VRE screening:** ICU, on admission and twice- weekly; haematology & oncology, renal, liver transplant wards, spinal ward, respiratory ward (ventilator support service) screened on admission and weekly<br>Biannual hospital-wide point-prevalence survey for *vanA* VRE and MRGN | ***vanA* VRE:** Transmission based precautions with single room with own bathroom, full gown, gloves<br>**ESBL-Ec and ESBL-Kp:** Transmission based precautions with single room, own bathroom, disposable apron |
| | **A2** | Subacute hospital, aged care and rehabilitation services | 150 | None | Biannual point-prevalence survey for *vanA* VRE and MRGN | |
| | **A3** | Subacute hospital, rehabilitation services | 60 | None | Biannual point-prevalence survey for *vanA* VRE and MRGN | |

**Table S1**. Overview of hospital network, ward specialties, screening strategies and infection prevention control measures. Table adapted from (6).

[a] Inpatient beds, excludes day cases, hospital-in-the-home and mental health.

| All target MDRO Clinical Samples | | | | | |
|---|---|---|---|---|---|
| | ICU | High-Consequence wards | High-Prevalence wards | Standard Risk wards | TOTAL |
| **Blood culture** | 2 | 13 | 3 | 15 | 33 |
| **Non-sterile site** | 1 | 5 | 0 | 4 | 10 |
| **Respiratory** | 3 | 2 | 0 | 0 | 5 |
| **Sterile site** | 3 | 4 | 0 | 2 | 9 |
| **Urine** | 4 | 29 | 14 | 61 | 108 |
| **TOTALS** | 13 | 53 | 17 | 82 | 165 |
| **Rate per 1000 patient separations** | 31.25 | 7.21 | 4.69 | 3.15 | 4.41 |

| Target MDRO Clinical Samples – blood cultures + sterile site samples only | | | | | |
|---|---|---|---|---|---|
| | ICU | High-Consequence wards | High-Prevalence wards | Standard Risk wards | TOTAL |
| **Blood culture** | 2 | 13 | 3 | 15 | 33 |
| **Sterile site** | 3 | 4 | 0 | 2 | 9 |
| **TOTALS** | 5 | 17 | 3 | 17 | 42 |
| **Rate per 1000 patient separations** | 12.02 | 2.31 | 0.83 | 0.65 | |

**Table S2.** Distribution of MDRO clinical samples and incidence rates per 1000 patient separations.

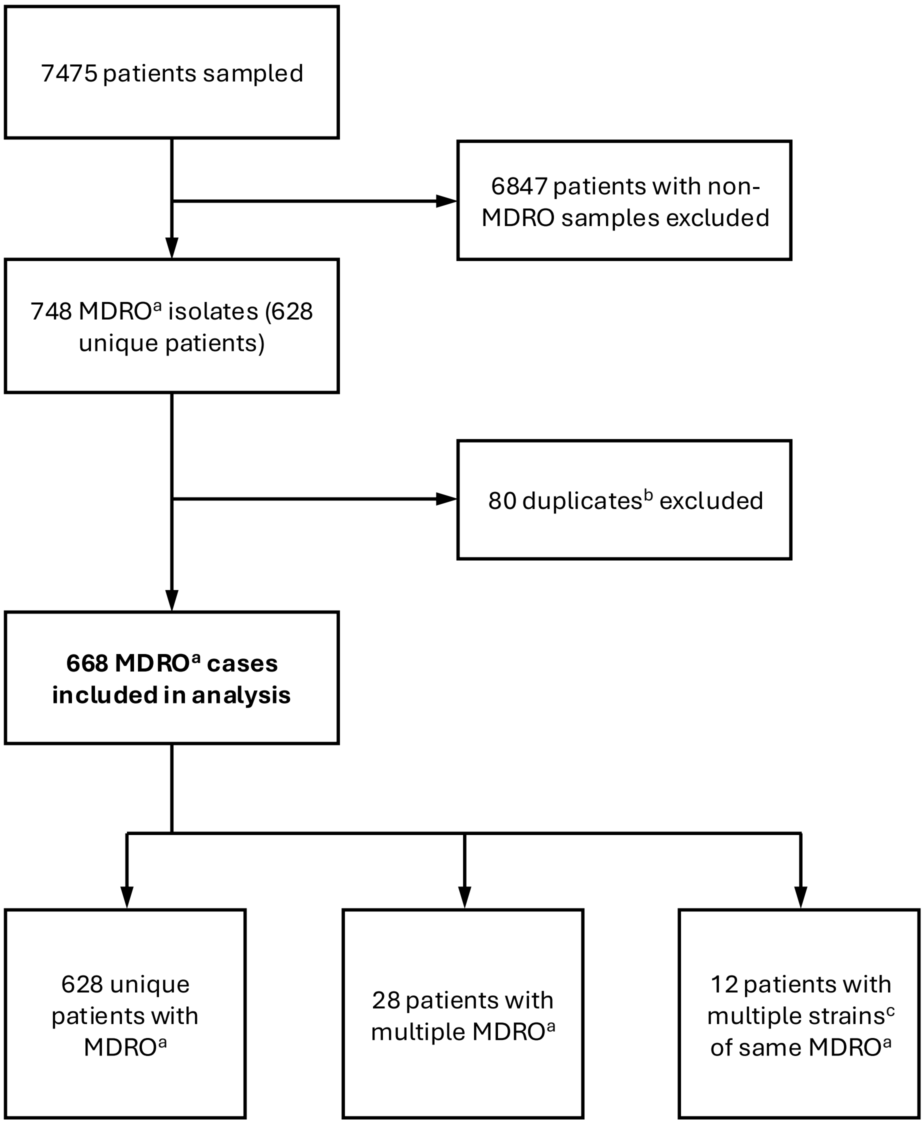


[a] Indicator MDRO comprised *vanA* VRE, ESBL *E. coli*, ESBL *K. pneumoniae*
[b] Duplicates = same MDRO strain in the same patient
[c] MDRO strains defined by multilocus sequence typing (MLST)
MDRO = multidrug-resistant organism

**Figure S1.** Number of patients, MDRO isolates and MDRO cases included in the study.

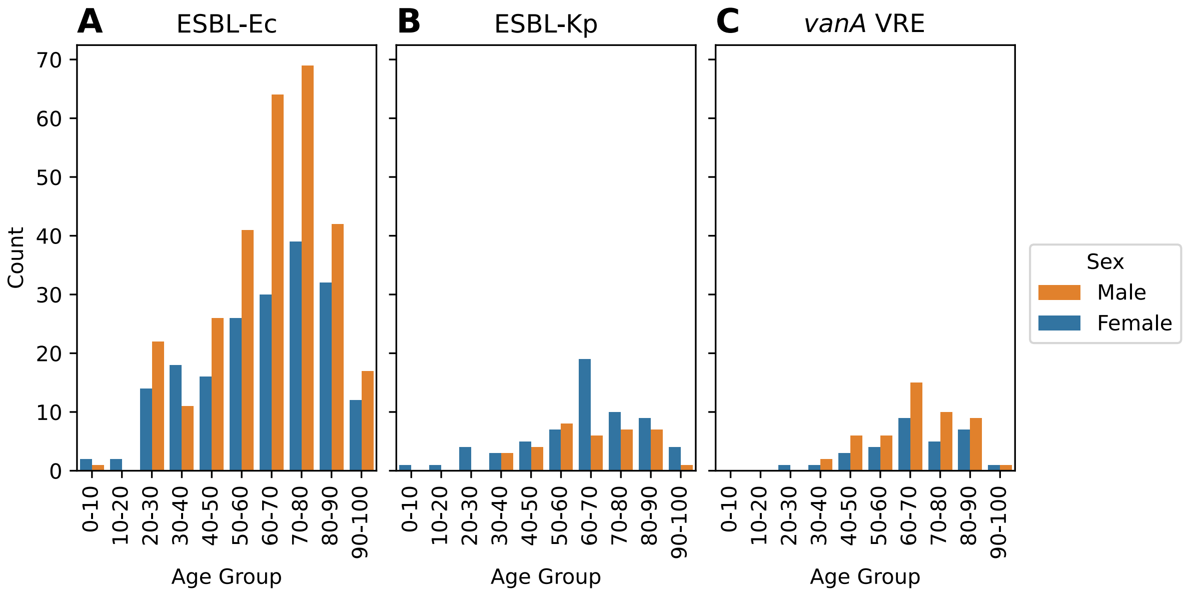


**Figure S2**. Frequency distributions of samples grouped by patient age. Bars are coloured by patient sex (male in orange, female in blue). Count of ESBL-Ec samples (A), count of ESBL-Kp samples (B) and count of *vanA* VRE samples (C).

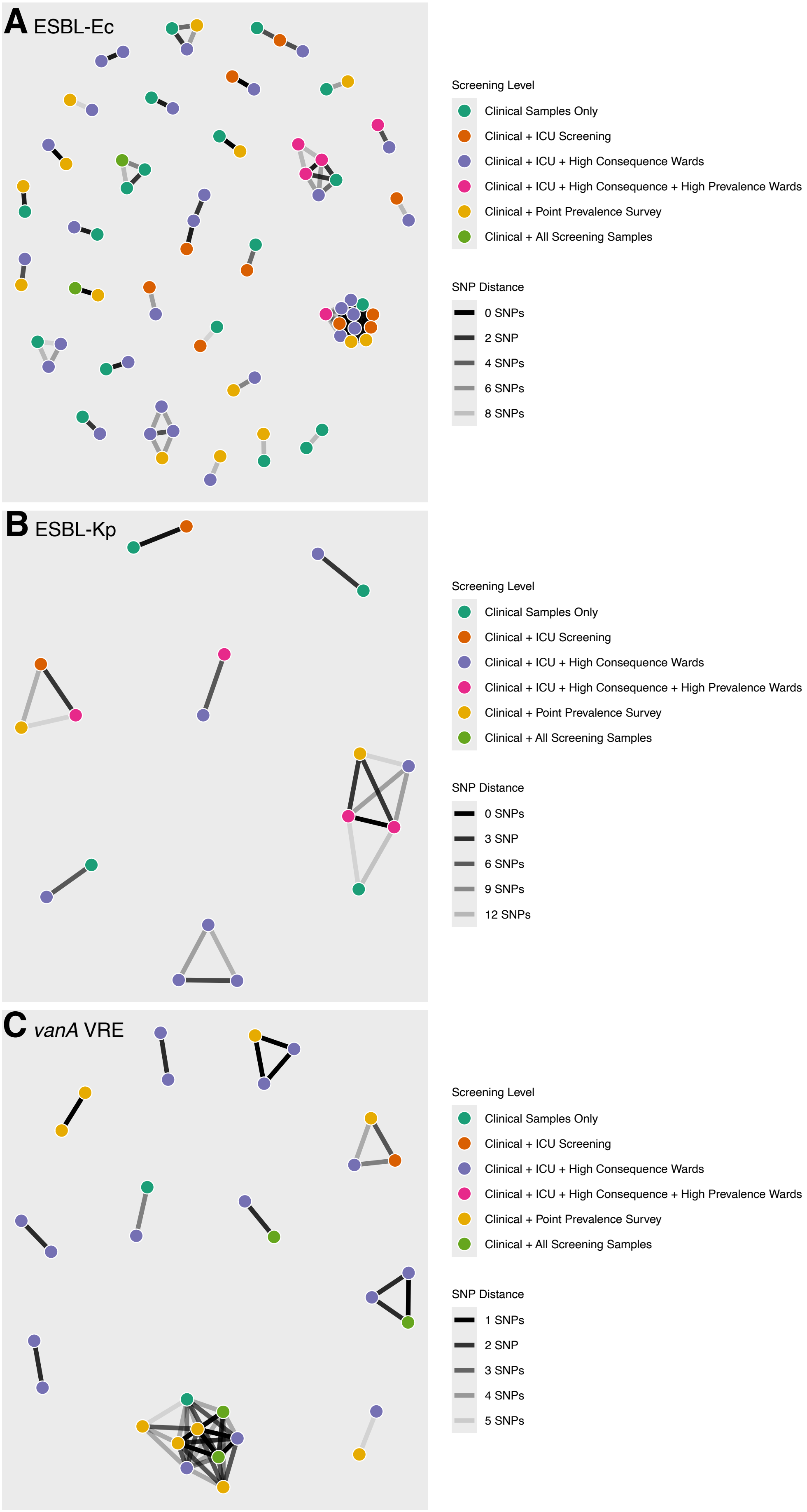
A ESBL-Ec
B ESBL-Kp
C vanA VRE
Screening Level
Clinical Samples Only
Clinical + ICU Screening
Clinical + ICU + High Consequence Wards
Clinical + ICU + High Consequence + High Prevalence Wards
Clinical + Point Prevalence Survey
Clinical + All Screening Samples
SNP Distance
0 SNPs
2 SNP
4 SNPs
6 SNPs
8 SNPs
Screening Level
Clinical Samples Only
Clinical + ICU Screening
Clinical + ICU + High Consequence Wards
Clinical + ICU + High Consequence + High Prevalence Wards
Clinical + Point Prevalence Survey
Clinical + All Screening Samples
SNP Distance
0 SNPs
3 SNP
6 SNPs
9 SNPs
12 SNPs
Screening Level
Clinical Samples Only
Clinical + ICU Screening
Clinical + ICU + High Consequence Wards
Clinical + ICU + High Consequence + High Prevalence Wards
Clinical + Point Prevalence Survey
Clinical + All Screening Samples
SNP Distance
1 SNPs
2 SNP
3 SNPs
4 SNPs
5 SNPs

**Figure S3**. Network visualizations of the full dataset. Coloured nodes represent cases for ESBL-Ec (A), ESBL-Kp (B) and *vanA* VRE (C). Nodes are coloured by screening tier. Black lines connecting nodes represent SNP distances below the species-specific threshold. Connecting line transparency represents the SNP distance between nodes with a fainter line corresponding to a greater SNP distance. Only cases that are linked to another case are shown. The full dataset (shown here) has duplicate patient samples excluded with only the first collected sample kept. Patient de-duplication occurred at each analysis level, hence patients who had a screening sample that preceded a clinical sample would have the clinical sample included for tier 1 but would be replaced by the screening sample at subsequent analysis tiers.

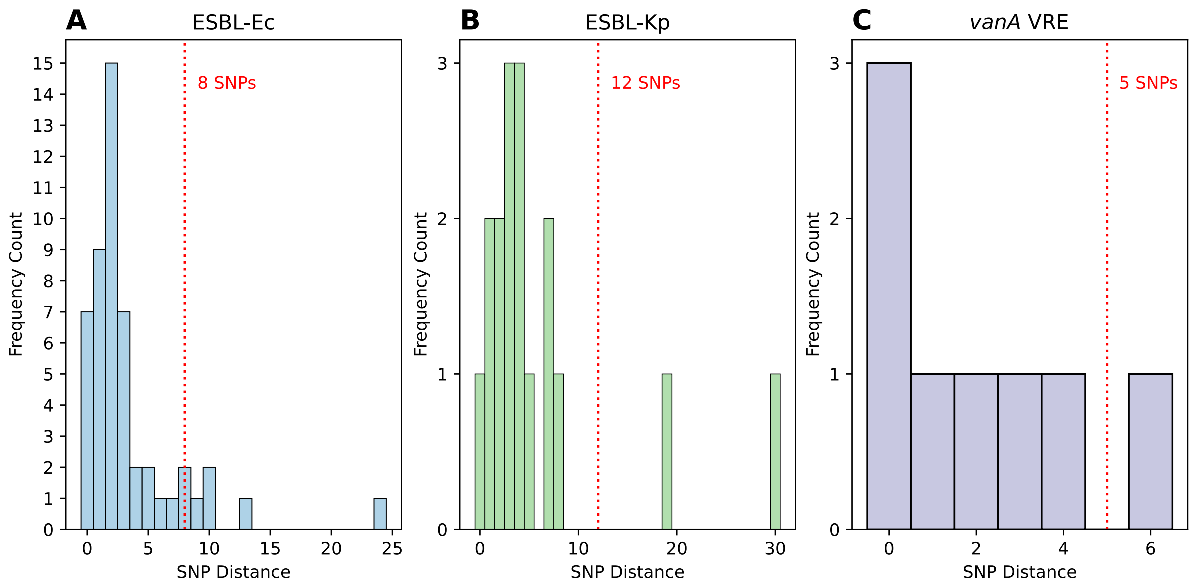


**Figure S4**. Frequency histograms of within patient SNP distances. Red line indicates the 90th percentile of the distribution rounded to nearest whole number. Distribution for ESBL-Ec (A), distribution for ESBL-Kp (B) and distribution for *vanA* VRE (C).
